# Association study of *DNAJC13, UCHL1, HTRA2, GIGYF2* and *EIF4G1* with Parkinson’s disease

**DOI:** 10.1101/2020.06.26.20141176

**Authors:** Prabhjyot Saini, Uladzislau Rudakou, Eric Yu, Jennifer A. Ruskey, Farnaz Asayesh, Sandra B. Laurent, Dan Spiegelman, Stanley Fahn, Cheryl Waters, Oury Monchi, Yves Dauvilliers, Nicolas Dupré, Lior Greenbaum, Sharon Hassin-Baer, Alberto J. Espay, Guy A. Rouleau, Roy N. Alcalay, Edward A. Fon, Ronald B. Postuma, Ziv Gan-Or

**Affiliations:** Montreal Neurological Institute, McGill University, Montréal, QC, H3A 1A1, Canada; Department of Human Genetics, McGill University, Montréal, QC, H3A 1A1, Canada; Department of Neurology and neurosurgery, McGill University, Montréal, QC, H3A 0G4, Canada, Canada; Department of Neurology, College of Physicians and Surgeons, Columbia University Medical Center, 710 West 168th Street, New York, NY, 10032-3784 USA; Department of Clinical Neurosciences and Department of Radiology, University of Calgary, 2500 University Drive NW Calgary, Alberta, T2N 1N4, Canada; Cumming School of Medicine, Hotchkiss Brain Institute, Calgary, 3330 Hospital Drive NW Calgary, Alberta, T2N 4N1 Canada; National Reference Center for Narcolepsy, Sleep Unit, Department of Neurology, Gui-de-Chauliac Hospital, CHU Montpellier, University of Montpellier, Inserm U1061, Montpellier, France; Division of Neurosciences, CHU de Québec, Université Laval, 2705 Laurier Boulevard, Quebec City, Quebec G1V 4G2, Canada; Department of Medicine, Faculty of Medicine, Université Laval, 1050 Avenue de la Médecine, Québec, QC G1V 0A6, Canada; Sackler Faculty of Medicine, Tel Aviv University, Tel Aviv, Israel; Danek Gertner Institute of Human Genetics, Sheba Medical Center, Tel Hashomer, Israel; Joseph Sagol Neuroscience Center, Sheba Medical Center, Tel Hashomer, Israel; The Movement Disorders Institute, Department of Neurology, Sheba Medical Center, Tel Hashomer, Israel; Gardner Family Center for Parkinson’s Disease and Movement Disorders, Department of Neurology, University of Cincinnati, 3113 Bellevue Ave, Cincinnati, OH 45229, USA; Taub Institute for Research on Alzheimer’s Disease and the Aging Brain, College of Physicians and Surgeons, Columbia University Medical Center, 710 W 168th St 3, New York, NY 10032, USA

**Keywords:** Parkinson’s Disease, DNAJC13, UCHL1, HTRA2, GIGYF2, EIF4G1, Ashkenazi Jewish, French-Canadian, SKAT-O

## Abstract

Rare mutations in genes originally discovered in multi-generational families have been associated with increased risk of Parkinson’s Disease (PD). The involvement of rare variants in *DNAJC13, UCHL1, HTRA2, GIGYF2* and *EIF4G1* loci have been poorly studied or produced conflicting results across cohorts. However, they are still being often referred to as “PD-genes” and used in different models. To further elucidate the role of these five genes in PD, we fully sequenced them using molecular inversion probes in 2,408 PD patients and 3,444 controls from 3 different cohorts. A total of 788 rare variants were identified across the five genes and three cohorts. Burden analyses and optimized sequence Kernel association tests revealed no significant association between any of the genes and PD after correction for multiple comparisons. Our results do not support an association of the five tested genes with PD. Combined with previous studies, it is unlikely that any of these genes plays an important role in PD. Their designation as “*PARK*” genes should be reconsidered.

## 1.0 Introduction

Parkinson’s disease (PD) is clinically characterized by progressive movement disability, mainly bradykinesia, tremor and muscle rigidity, often accompanied by other motor and non-motor symptoms (Jankovic, 2008). Common genetic variants of PD have largely been discovered through genome-wide association studies (GWAS), and to date 92 independent risk variants in 80 loci have been identified (Foo et al., 2020; Nalls et al., 2019). Other types of studies, including linkage and next generation sequencing studies in families with multiple affected family members with PD or other forms of Parkinsonism led to the identification of familial PD-associated genes, many of them have received the alias *PARK* (e.g. *PARK1, PARK2*, etc.) (Deng et al., 2018).

Five suggested PD-causing genes identified through family studies and cohorts, *UCHL1* (Lincoln et al., 1999), *GIGYF2* (Lautier et al., 2008), *HTRA2* (Strauss et al., 2005), *EIF4G1* (Chartier-Harlin et al., 2011), and *DNAJC13* (Vilariño-Güell et al., 2013), (also termed *PARK5, PARK11, PARK13, PARK18* and *PARK21*, respectively) have been under scrutiny regarding their association with PD. Their discovery shares several similarities, including the presumed role of loss-of-function, missense mutations and dominant heritability (Nalls et al., 2019). Subsequent studies, in many cases, failed to replicate the association of these genes with PD (Bartonikova et al., 2018; Gagliardi et al., 2018; Kruger et al., 2011; Nuytemans et al., 2013; Sun et al., 2014). However, ongoing studies, including those using cellular and animal models, continue to refer to these genes as PD-associated genes, and to invoke their function in PD-related mechanisms (Chen et al., 2018; Tran et al., 2018).

The primary goal of the present study is to use large cohorts of PD patients (n=2,382) and controls (n=3,411) to further examine the potential role of rare variants in these genes in PD.

## 2.0 Materials & Methods

### 2.1 Populations

Full sequencing of *UCHL1, HTRA2, GIGYF2, EIF4G1* and *DNAJC13* was performed in 2,408 PD patients and 3,444 controls from three different cohorts: a) McGill cohort, including 855 patients and 2,441 controls from Quebec, Canada and Montpellier, France, all of European origin, b) Columbia cohort, including 963 patients and 508 controls from New York, Mainly of European or Ashkenazi-Jewish (AJ) ancestry, and c) Sheba cohort, including 590 patients and 495 controls of AJ ancestry collected at Sheba Medical Center, Israel. Demographic details on these cohort can be found in Supplementary Table S1. All subjects were consecutively recruited, and patients were diagnosed by a movement disorders specialist according to the UK Brain Bank Criteria (Hughes et al., 1992) or the MDS criteria (Postuma et al., 2015). All patients and controls signed informed consent at enrollment and the study protocols were approved by the institutional review.

### 2.2 Genetic analysis

The entire coding regions of the five genes, including 5’ and 3’ untranslated regions (UTRs) and exon-intron boundaries, were captured using molecular inversion probes (MIPs), followed by sequencing as previously described (Ross et al., 2016). Supplementary Table S2 includes all the MIPs used to capture and sequence *GIGYF2, HTRA2, DNAJC13, UCHL1, EF4G1*, and the full protocol is available upon request. The library was sequenced using Illumina HiSeq 2500/4000 platform at the McGill University and Genome Quebec Innovation Centre. Reads were mapped to the human reference genome (hg19) with Burrows-Wheeler Aligner (Li and Durbin, 2009). Alignment, quality control and variant calling was performed with Genome Analysis Toolkit (GATK, v3.8) (McKenna et al., 2010) and PLINK software v1.9 (Purcell et al., 2007) and ANNOVAR was used for annotation (Wang, K. et al., 2010). Minor allele frequency from European and Ashkenazi Jewish ancestries were extracted from the public database Genome Aggregation Database (GnomAD) (Lek et al., 2016). The variants were filtered based on a minimum read depth ≥30X, a genotype quality (GQ) ≥30, genotyping rate cut-off for individuals was 90%, missingness difference between patients and controls was set at *p*=0.05 and adjusted by Bonferroni correction, proportion of the reads with alternative alleles ≥ 25%, and deviation from Hardy-Weinberg equilibrium was set at *p*=0.001.

### 2.3 Statistical Analyses

Burden and optimized Sequence Kernel Association Test (SKAT-O, R package) (Lee et al., 2012) were used to analyze the joint effects of rare variants (minor allele frequency, MAF ≤0.01). These analyses were performed on five groups of variants: a) all rare variant, b) functional variants, defined as all nonsynonymous, splice-site, frameshift and stop variants, c) loss of function variants, defined as splice-site, frameshift and stop variants, d) nonsynonymous variants and e) variants with Combined Annotation Dependent Depletion (CADD) score of >12.37, which is the threshold for the top 2% of predicted most deleterious variants. We repeated the analysis twice, at coverage of >30X and at coverage of >50X, to ensure high quality and confidence in the variant calls. Bonferroni correction for multiple comparisons considered the number of genes (five), number of cohorts (three), number of depth coverage (two) and number of statistical tests (two), which set the threshold for statistical significance at p<0.00083.

## 3.0 Results

### 3.1 Quality control and variants identified

The average coverage of the five genes range between 237X – 1592X in the different cohorts, and the percentages of read depth of the targeted genes were 93.07% and 88.76% of the bases covered by at least 20X and 50X respectively, and the coverage of each gene was nearly identical in the three cohorts (coverage depth and average coverage for each gene in each cohort are detailed in Supplementary Table S3). We identified a total of 788 rare variants in these five genes. The distribution of rare variants across the coding region and functional domains is available across cohorts and sequencing depth (Supplementary Table S4).

### 3.2 Association of rare variants and risk of Parkinson’s disease

To further investigate whether rare variants in these five genes contribute to PD risk, we conducted a gene-based burden test and found no significant results within cohorts and depths across mutation type (Table 1). SKAT-O test for enrichment of all variant groups across the entire gene region in each cohort and sequencing depth suggested that *DNAJC13* variants were nominally enriched in PD patients in the Sheba cohort (p=0.0426) and were driven by nonsynonymous variants (p=0.0002) (Table 1). However, this association was driven by higher frequency of nonsynonymous variants in controls (1.9%) than in PD patients (1.1%, Supplementary Table S5), demonstrating that nonsynonymous variants in this gene are not associated with higher risk of PD. In all other four genes there were no statistically significant associations in any of the cohorts tested (Table 1).

**Table 1:** SKAT-O and burden analysis results for each gene, across cohort and coverage depth for mutation type

| Depth of coverage | Gene | All rare |  | Rare CADD |  | Rare functional |  | Rare nonsynonymous |  | Rare loss-of-function |  |
| --- | --- | --- | --- | --- | --- | --- | --- | --- | --- | --- | --- |
|  |  | SKAT-O | SKAT Burden | SKAT-O | SKAT Burden | SKAT-O | SKAT Burden | SKAT-O | SKAT Burden | SKAT-O | SKAT Burden |
| 30x | McGill |  |  |  |  |  |  |  |  |  |  |
|  | *DNAJC13* | 0.7274 | 0.4935 | 0.9052 | 1.0000 | 0.8449 | 0.6467 | 0.8987 | 0.8001 | NV | NV |
|  | *EIF4G1* | 0.7330 | 0.8183 | 0.6368 | 0.9859 | 0.5659 | 0.8356 | 0.8719 | 0.6904 | 0.3153 | 1.0000 |
|  | *GIGYF2* | 0.2543 | 0.2358 | 0.2366 | 0.4767 | 0.1037 | 0.0750 | 0.3201 | 0.6587 | 0.8845 | 0.8539 |
|  | *HTRA2* | 0.5922 | 0.3411 | 0.7980 | 0.7980 | 0.2923 | 0.5129 | 0.2228 | 0.2228 | 0.6114 | 0.6114 |
|  | *UCHL1* | 0.8267 | 1.0000 | 0.7288 | 0.3405 | 0.5023 | 1.0000 | 0.4795 | 0.3130 | NV | NV |
|  | Columbia |  |  |  |  |  |  |  |  |  |  |
|  | *DNAJC13* | 0.7085 | 0.4927 | 0.3437 | 0.2061 | 0.5000 | 0.3173 | 0.5935 | 0.3928 | NV | NV |
|  | *EIF4G1* | 0.5109 | 0.3411 | 0.9826 | 0.8615 | 0.3835 | 0.2392 | 0.6344 | 0.4288 | 0.4060 | 0.7585 |
|  | *GIGYF2* | 0.7827 | 0.5762 | 0.6380 | 0.5549 | 0.7934 | 0.6399 | 0.8887 | 0.7576 | 0.5551 | 0.5836 |
|  | *HTRA2* | 0.2764 | 0.3233 | 0.3438 | 0.7742 | 0.8353 | 0.8353 | 0.3438 | 0.7742 | NV | NV |
|  | *UCHL1* | 0.7508 | 0.7154 | 0.9283 | 0.7706 | 0.8912 | 0.7413 | 0.7805 | 0.5467 | NV | NV |
|  | Sheba |  |  |  |  |  |  |  |  |  |  |
|  | *DNAJC13* | 0.0423 | 0.5694 | 0.0024 | 0.2642 | 0.0014 | 0.1520 | 0.0013 | 0.1174 | NV | NV |
|  | *EIF4G1* | 0.7028 | 0.4890 | 0.5436 | 0.6780 | 0.6174 | 0.5828 | 0.6353 | 0.6881 | 0.1026 | 0.1026 |
|  | *GIGYF2* | 0.2781 | 0.2030 | 0.8367 | 0.7090 | 0.1977 | 0.2324 | 0.6536 | 0.4732 | 0.1654 | 0.1593 |
|  | *HTRA2* | 0.0661 | 0.0803 | 0.0463 | 0.0463 | 0.2641 | 0.1382 | 0.0463 | 0.0463 | NV | NV |
|  | *UCHL1* | 0.4786 | 0.3435 | 0.8450 | 0.8450 | 0.5739 | 0.3347 | 0.9178 | 0.7945 | 1.0000 | 1.0000 |
| 50x | McGill |  |  |  |  |  |  |  |  |  |  |
|  | *DNAJC13* | 0.6015 | 0.5437 | 0.3990 | 1.0000 | 0.4100 | 0.8328 | 0.4158 | 0.8600 | NV | NV |
|  | *EIF4G1* | 0.4384 | 1.0000 | 0.8636 | 0.8636 | 0.2985 | 1.0000 | 0.9448 | 0.8344 | 0.2413 | 0.8701 |
|  | *GIGYF2* | 0.3312 | 0.3512 | 0.1292 | 0.9995 | 0.0933 | 0.1360 | 0.1628 | 0.7996 | 0.8448 | 0.8448 |
|  | *HTRA2* | NV | NV | NV | NV | NV | NV | NV | NV | NV | NV |
|  | *UCHL1* | 0.4345 | 0.2882 | 0.7152 | 0.7152 | 0.2815 | 0.1934 | 0.2815 | 0.1934 | NV | NV |
|  | Columbia |  |  |  |  |  |  |  |  |  |  |
|  | *DNAJC13* | 0.6194 | 0.4114 | 0.3587 | 0.2184 | 0.3861 | 0.2361 | 0.5021 | 0.3224 | NV | NV |
|  | *EIF4G1* | 0.5684 | 0.5900 | NV | NV | 0.3289 | 0.9888 | NV | NV | 0.3314 | 0.3314 |
|  | *GIGYF2* | 0.3701 | 0.2280 | 0.4528 | 0.2644 | 0.3080 | 0.1814 | 0.6180 | 0.4336 | 0.2021 | 0.1348 |
|  | *HTRA2* | NV | NV | NV | NV | NV | NV | NV | NV | NV | NV |
|  | *UCHL1* | 0.5247 | 0.6941 | 0.4676 | 0.3779 | 0.6953 | 0.9221 | 0.6953 | 0.9221 | NV | NV |
|  | Sheba |  |  |  |  |  |  |  |  |  |  |
|  | *DNAJC13* | 0.0426 | 0.1512 | 0.0002 | 0.1323 | 0.0005 | 0.1389 | 0.0002 | 0.1323 | NV | NV |
|  | *EIF4G1* | 0.6150 | 0.3688 | 0.7538 | 0.7538 | 0.7538 | 0.7538 | 0.7538 | 0.7538 | NV | NV |
|  | *GIGYF2* | 0.6256 | 0.4155 | 0.6067 | 0.4443 | 0.5098 | 0.3352 | 0.7803 | 0.6084 | NV | NV |
|  | *HTRA2* | NV | NV | NV | NV | NV | NV | NV | NV | NV | NV |
|  | *UCHL1* | 0.5455 | 0.3953 | 0.8472 | 0.8472 | 0.9166 | 0.7915 | 0.9166 | 0.7915 | NV | NV |

Abbreviations: SKAT: Sequence Kernel Association Test; SKAT-O: Optimized Sequence Kernel Association Test; CADD: Combined Annotation Dependent Depletion; NV: No Variant

## 4.0 Discussion

Our study, which included full sequencing of *UCHL1, GIGYF2, HTRA2, DNAJC13* and *EIF4G1* in 3 cohorts, identified 788 rare variants (MAF <0.01) that are nonsynonymous, loss-of-function, pathogenic as predicted by CADD scores, or affect splicing in coding regions of the gene and splice sites. Our results do not support a role for any of these genes in PD.

Of the five genes, *DNAJC13* in the Sheba cohort showed nominal association with controls driven by nonsynonymous variants with higher frequency in controls compared to PD cases. A previously reported variant, p.L1207W (Ross et al., 2016), was detected in all 3 cohorts, but had similar frequencies between cases and controls (Columbia), greater frequency in controls than cases (McGill) or were found exclusively in controls (Sheba, Supplementary Table S6-S8). The original *DNAJC13* association were described in a large-multi-incident family of Dutch-German-Russian-Mennonite ancestry that presented with autosomal dominant PD, where a p.N855S coding variant only partially co-segregated with disease status. Since the original description, several studies have reported other genetic variants in Italian cohorts (p.R903K) (Gagliardi et al., 2018) and Taiwanese cohorts (p.G394V and p.R1382H), yet the pathogenicity of these variants has not been confirmed. In addition, other previously described variants did not segregate by disease (p.R1516H and p.L2170W) (Ross et al., 2016), and in cohorts with sparse number of probands with rare variants in *DNAJC13* (Gagliardi et al., 2018; Lin et al., 2019). No other study was able to confirm the pathogenicity of the *DNAJC13* p.N855S variant. Moreover, in the same Dutch-German-Russian-Mennonite family, another gene, *TMEM230*, has been reported to be associated with PD instead of *DNAJC13*. Similar to *DNAJC13, TMEM230* had imperfect disease segregation within the family (Deng et al., 2016). These inconsistent results, together with the negative results in the current study and previous studies (Foo et al., 2014; Lorenzo-Betancor et al., 2015) call into question a role for *DNAJC13* in PD.

The role of *HTRA2* in PD was initially suggested as two missense variants (p.G399S and p.A141S) were reported to be associated with PD (Strauss et al., 2005). Subsequently, the p.G399S variant co-segregated with PD and essential tremor (ET) in a large Turkish family (Unal Gulsuner et al., 2014), providing further evidence of its potential role in PD. Additional variants were reported in Belgian (p.R404W) (Bogaerts et al., 2008), Chinese (IVS5+29T>A) (Wang, C.Y. et al., 2011), and Taiwanese (p.P143A) PD patients (Lin et al., 2011), yet without evidence for pathogenicity. Evidence for the p.G399S variant remains elusive even after several large-scale studies with PD in various populations worldwide (Kruger et al., 2011; Ross et al., 2008; Simon-Sanchez and Singleton, 2008). Moreover, the p.G399S mutations has been associated with ET, yet in subsequent studies of ET and PD, there was no significant evidence of an association of this variant with PD (He et al., 2017). Similar to *DNAJC13*, there are insufficient data to conclude that this gene is associated with PD.

Since the first description of the association of *EIF4G1* variants (p.R1205H and p.A502V) with PD (Chartier-Harlin et al., 2011), numerous subsequent studies could not conclude that *EIF4G1* variants caused or associated with PD (Nuytemans et al., 2013; Schulte et al., 2012; Siitonen et al., 2013; Tucci et al., 2012). It is possible that the p.R1205H variant is not highly penetrant, as it was found in the original study (Chartier-Harlin et al., 2011) and in another family where it co-segregated with PD (Nuytemans et al., 2013); however, it was also reported in three controls (Schulte et al., 2012). Ethnic differences could account for the lack of findings as these variants were not found in several Asian ethnicities (Chen et al., 2013; Li et al., 2013; Nishioka et al., 2014; Sudhaman et al., 2013; Zhao et al., 2013). Interestingly, in the present study, the p.R1205H variant was found exclusively in controls in the McGill and Columbia cohorts. Given the multiple negative results and the presence of the p.R1205H in multiple controls, *EIF4G1* is unlikely to play a role in PD.

*GIGYF2* was initially reported in a cohort composed of Italian and French PD patients (Lautier et al., 2008), yet subsequent studies in Portuguese and US cohorts did not find evidence of association with PD (Bras et al., 2009). Numerous additional studies, including in a different Italian cohort and multiple other ethnicities, also failed to identify an association of *GIGYF2* variants with PD (Bartonikova et al., 2018; Bonetti et al., 2009; Di Fonzo et al., 2009; Guo et al., 2009; Huo et al., 2017; Lesage et al., 2010; Li et al., 2010; Meeus et al., 2011; Nichols et al., 2009; Samaranch et al., 2010; Tan et al., 2009; Tan and Schapira, 2010; Tian et al., 2012; Vilarino-Guell et al., 2009; Wang, L. et al., 2010; Wang, L. et al., 2011; Yang et al., 2019; Zhang et al., 2015; Zhang et al., 2009; Zimprich et al., 2009). It is therefore very unlikely that *GIGYF2* is associated with PD.

The association of *UCHL1* was initially reported with the common p.S18Y variant (Lincoln et al., 1999), and while several studies initially supported this association (Maraganore et al., 2004; Ragland et al., 2009) other studies have failed to demonstrate that any *UCHL1* variants are associated with increased risk of PD (Elbaz et al., 2003; Healy et al., 2006; Levecque et al., 2001; Mellick and Silburn, 2000; Momose et al., 2002; Satoh and Kuroda, 2001; Savettieri et al., 2001; Wang et al., 2002; Wintermeyer et al., 2000; Zhang et al., 2000). Since the *UCHL1* p.S18Y is common, it should have been discovered in the large European and Asian GWASs (Foo et al., 2020; Nalls et al., 2019) yet it has not. In the present study, rare variants in *UCHL1* also did not demonstrate any association with PD. Given with the lack of effect of common variants, *UCHL1* is unlikely to be involved in PD.

Our study has several limitations, including the lack of age and sex matching between patients and controls. However, in the Columbia cohort the controls were older than the patients, whereas in the other two cohorts the controls were younger, yet no significant associations were found in any, suggesting that age did not affect the current results. In addition, it is possible that control subjects develop PD in the future, specifically in those cases where variants were found exclusively in controls. Since the risk of PD is about 1-2% in the control population, the effect on the results would likely be minor, even if a few individuals did develop PD. Lastly, despite being a large study, we cannot completely rule out that very rare variants or more common variants with a very small effect on risk could be involved in PD in one or more of five tested genes. The fact that many large studies, including GWASs, genotyping and sequencing studies have so far mostly failed to detect associations, makes this possibility seem unlikely.

Altogether, the present study and the survey of existing literature strongly indicate a lack of association of PD with *UCHL1, HTRA2, GIGYF2, EIF4G1* and *DNAJC13*. The *PARK* aliases that these genes have received may lead to confusion and continue to inspire basic and clinical research as PD models. As the evidence suggests these genes are not associated with PD, we recommend removing the *PARK* designation off these genes.

## Data Availability

Anonymized data is available upon request by any qualified investigator.

## Acknowledgements

We thank the patients and control subjects for their participation in this study. This work was financially supported by the Michael J. Fox Foundation, the Canadian Consortium on Neurodegeneration in Aging (CCNA), the Canada First Research Excellence Fund (CFREF), awarded to McGill University for the Healthy Brains for Healthy Lives (HBHL) program and Parkinson Canada. The Columbia University cohort is supported by the Parkinson’s Foundation, the National Institutes of Health [K02NS080915, and UL1 TR000040] and the Brookdale Foundation. GAR holds a Canada Research Chair in Genetics of the Nervous System and the Wilder Penfield Chair in Neurosciences. EAF is supported by a Foundation Grant from the Canadian Institutes of Health Research (FDN grant – 154301) and a Canada Research Chair (Tier 1) in Parkinson Disease. ZGO is supported by the Fonds de recherche du Québec - Santé (FRQS) Chercheurs-boursiers award and Parkinson Quebec, and by the Young Investigator Award by Parkinson Canada. The access to part of the participants for this research has been made possible thanks to the Quebec Parkinson’s Network (http://rpq-qpn.ca/en/). We thank Daniel Rochefort, Helene Catoire, Clotilde Degroot and Vessela Zaharieva for their assistance.

## Conflict of Interest

SF received consulting fees/honoraria for board membership (unrelated to the current study) from Retrophin Inc., Sun Pharma Advanced Research Co., LTD and Kashiv Pharma. CW received consulting fees/honoraria (unrelated to the current study) from US World Meds, Acadia, Lundbeck, Cynapsus, Acorda. ND received consultancy fees (unrelated to the current work) from Actelion Pharmaceuticals. SHB received consulting fees from Actelion Pharmaceuticals Ltd., Abbvie Israel, Robotico Ltd., Medtronic Israel, Medison Pharma Israel (unrelated to the current study). AJE received grant support from the NIH and the Michael J Fox Foundation; personal compensation as a consultant/scientific advisory board member for Abbvie, Neuroderm, Neurocrine, Amneal, Adamas, Acadia, Acorda, InTrance, Sunovion, Lundbeck, and USWorldMeds; publishing royalties from Lippincott Williams & Wilkins, Cambridge University Press, and Springer; and honoraria from USWorldMeds, Acadia, and Sunovion. RNA received consultation fees (unrelated to the current study) from Biogen, Denali, Genzyme/Sanofi and Roche. EAF Received consulting fees (unrelated to the current study) from Inception Sciences. ZGO received consulting fees (unrelated to the current study) from Denali, Inception Sciences (now Ventus), Idorsia, Lysosomal Therapeutics Inc., Prevail Therapeutics, Deerfield, Neuron23 and Handl Therapeutics. All other authors report no conflict of interest.

## Notes

### Author Declarations

All patients and controls signed informed consent at enrollment and the study protocols were approved by the institutional review board of McGill University.

